# Remarks on pooling Coronavirus tests

**DOI:** 10.1101/2020.06.08.20125781

**Authors:** Alexander Pikovski, Kajetan Bentele

**Affiliations:** self-employed; no institution

## Abstract

Diagnostic testing for the novel Coronavirus is an important tool to fight the Covid-19 pandemic. However, testing capacities are limited. A modified testing protocol, whereby a number of probes are “pooled” (that is, grouped), is known to increase the capacity for testing. Here, we model pooled testing with a double-average model, which we think to be close to reality for Covid-19 testing. The optimal pool size and the effect of test errors are considered. Results show that the best pool size is three to five, under reasonable assumptions. Pool testing even reduces the number of false positives in the absence of dilution effects.

## Introduction / Main

The fast global spread of the novel Coronavirus is responsible for current pandemic, known as Covid-19. Diagnostic tests for the novel Coronavirus (nCov) are being employed worldwide. It is self-evident that increasing the number of diagnostic tests is important to combat the epidemic.

Widely used tests for Covid-19 detect the novel Coronavirus RNA genome, using the real-time polymerase chain reaction method. The capacities for such tests, however, are limited by the use reactants, machine time, lab personnel time, and overall logistics. Remarkably, testing capacities can be increased by a modification of the testing protocol.

Combining probes from several individuals and testing them together reduces the total amount of tests needed. This method, known as pooling or group testing, has been known for a long time [1]. The main idea is: when probes from several people are mixed together (pooled) and tested, the test will report negative when everyone is healthy and positive when at least one person is infected.

This study deals with Dorfman’s classic two-stage protocol [1] (described below). More complicated procedures have been devised; however, and we shall not consider with them here.

The pooling method was validated for nCov tests by many laboratories [2, 3, 4, 5, 6, 7, 8, 9, 10, 11, 12, 13, 14, 15]. However, the following questions remained open, which we address in this paper. 1) What pool size should be used? 2) How is the procedure affected by test errors?

## Results

The two-stage procedure is as follows [1]. We denote the number of people in one pool, i.e. the pool size, by *M*. Probes of *M* people are each split in two parts. One part is mixed together with the others, while the other part is kept for later. The mixture is tested for nCov. Upon a negative result, all the *M* people are declared healthy. When the result is positive, go to the second stage. In the second stage, each of the *M* people is tested individually using the probes that were put aside previously; the final test result is that of this second stage.

### Pool size

Here we address the question: How many people should be pooled together?

We consider a “double-averaging” model (see Methods), where the prevalence is estimated by a minimal and maximal value. Then, the results are only weakly dependent on the precise values, for reasonable values. Taking the prevalence to be between 0 and 30%, we have the result that a pool size of four is optimal; see Fig. 1 for other ranges. Pool sizes of three and five give results which are close to optimal, see Fig. 2 for a comparison.

**Figure 1:**
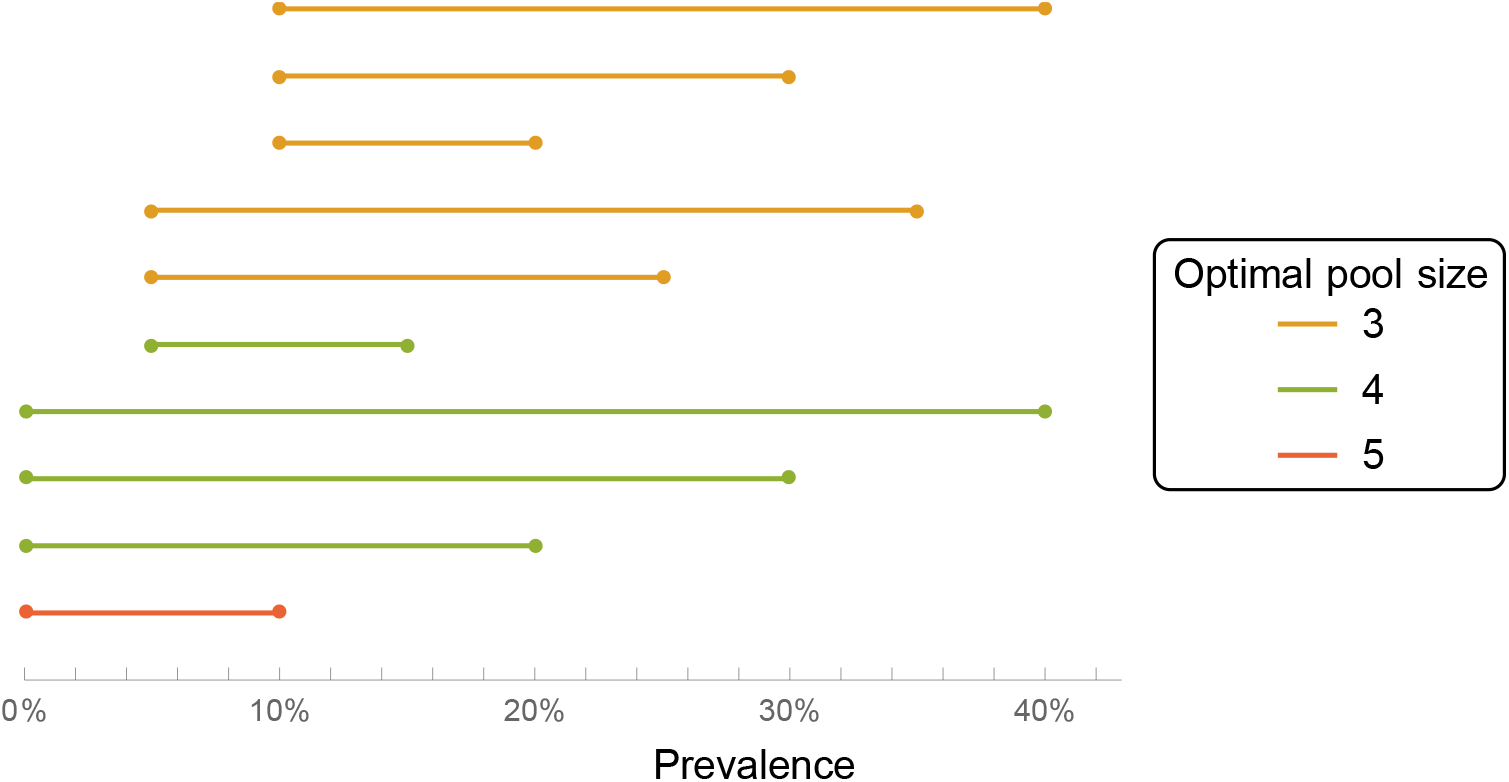
Optimal pool size (double-averging model), for different prevalence ranges.

**Figure 2:**
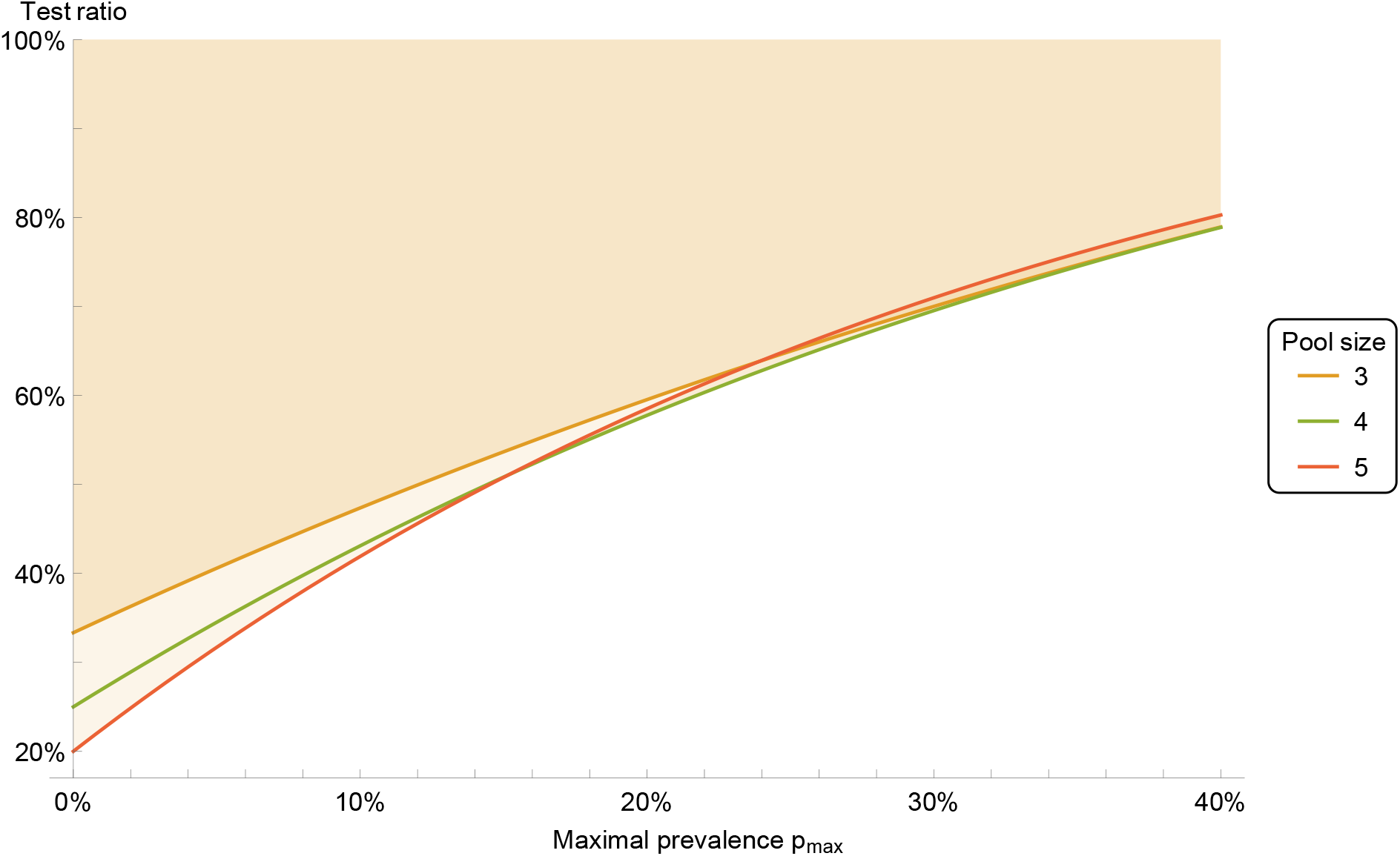
Test ratio (number of tests with pooling divided by number of tests without pooling), for prevalence range between 0 and *p*_max_. The colored area indicates improvement with respect to individual testing.

Therefore we recommend a pool size of four. Pool sizes of three or five can be used as well, if that is more convenient for practical reasons.

### Testing errors

Non-ideal tests produce erroneous results, which can be described by a false negative rate and a false positive rate, or equivalently, by giving the test sensitivity and specificity.

Two sources of errors arise in pooled tests. First, each test that is performed is inherently non-ideal and may give an error. These “single-test” errors are described by single-test sensitivity and specificity. Second, an error may arise if several probes are mixed together, due to dilution of the material from each individual. Such “dilution errors” are plausible for large pool sizes. We advocate small pool sizes (up to five), and at such small pool sizes no dilution errors have been found in lab studies [2, 14, 11, 16, 4]. Thus, for our analysis we assume no dilution errors.

The two-stage testing protocol leads to a modification of the overall error rates. For the pooled sensitivity, one obtains that the sensitivity of the pooled test is equal to the square of sensitivity of a single test. This means that the pooled sensitivity is reduced, and the number of false negatives increased. The specificity of the pooled test, on the other hand, is increased compared with the single test. (See Methods for details.) This means that the number of false positives is reduced. The reason for this increase of specificity is, that probes with infection are tested twice in pooling, but only once in individual tests. Figure 3 shows, as an example, the improvement of the false positive rate compared to single tests.

**Figure 3:**
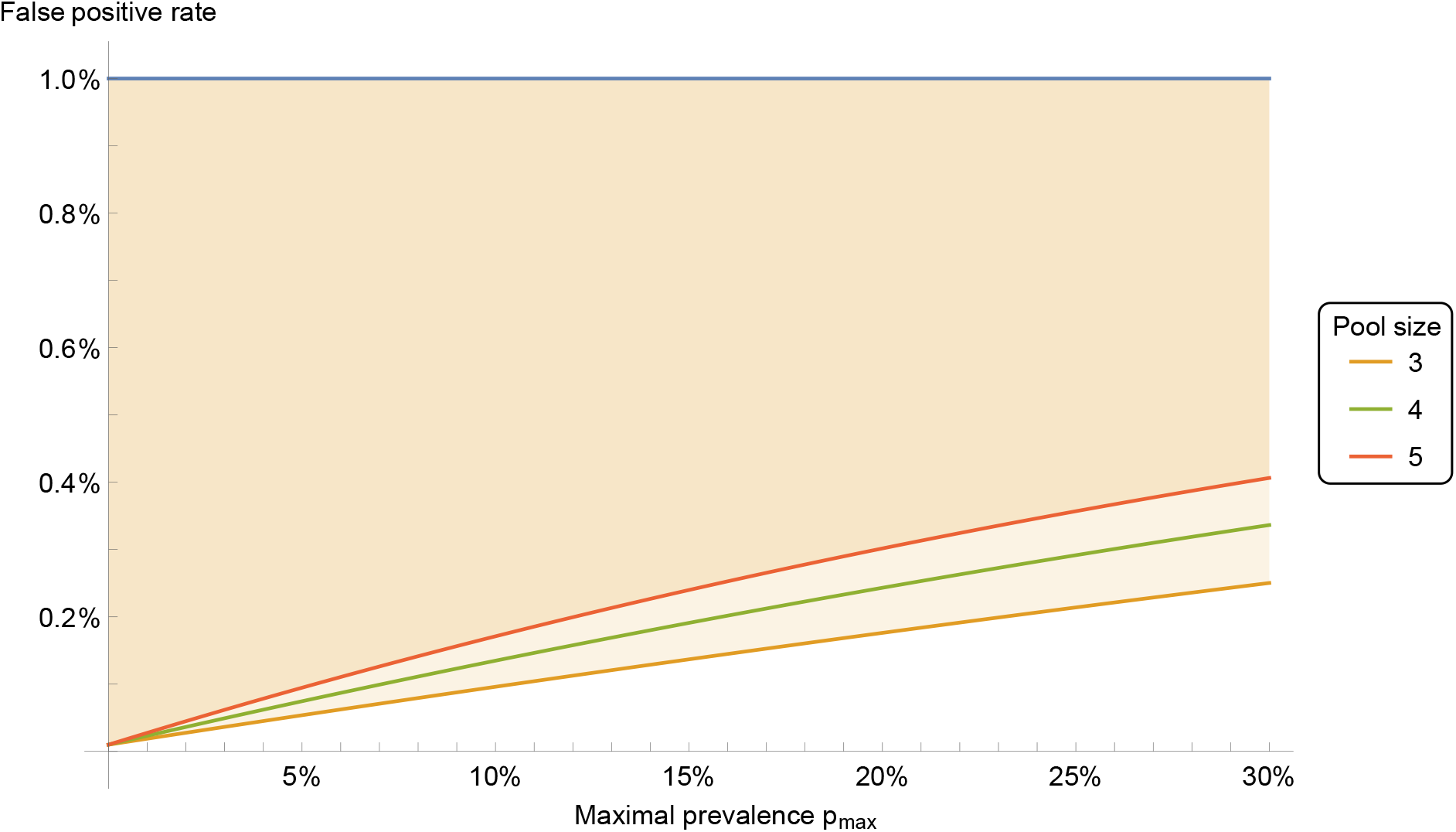
False positive ratio for pooled tests, for prevalence range between 0 and *p*_max_. The colored area indicates improvement with respect to individual testing. Here the individual test sensitivity is *s* = 90% and specificity *z* = 99%. Pool size is 4.

## Methods

When tests from *M* people are pooled, then the pool infection rate *p*′ (i.e. the probability that a pool is infected) is *p*′ = 1*−* (1 *−p*)^*M*^, where *p* is the prevalence in the statistical population. The number of tests performed during the two-stage pooling protocol (described above) will be denoted by *T*. Its expectation value for a statistical population of *N* individuals with prevalence *p* is *E*[*T*] = *N/M*+*N −N*(1*− p*)^*M*^, while the variance is Var[*T*] = *NM*((1*− p*)^*M*^*−* (1 *−p*)^2*M*^). It is convenient to present the results in terms of the test ratio *t* = *T/N*, which is the ratio of the number of tests performed with pooling to the number of tests without pooling. Smaller ratios indicate better performance of pooling. A plot of the test ratio as a function of the prevalence, with error bars due to the variance, is shown in Fig. 4.

**Figure 4:**
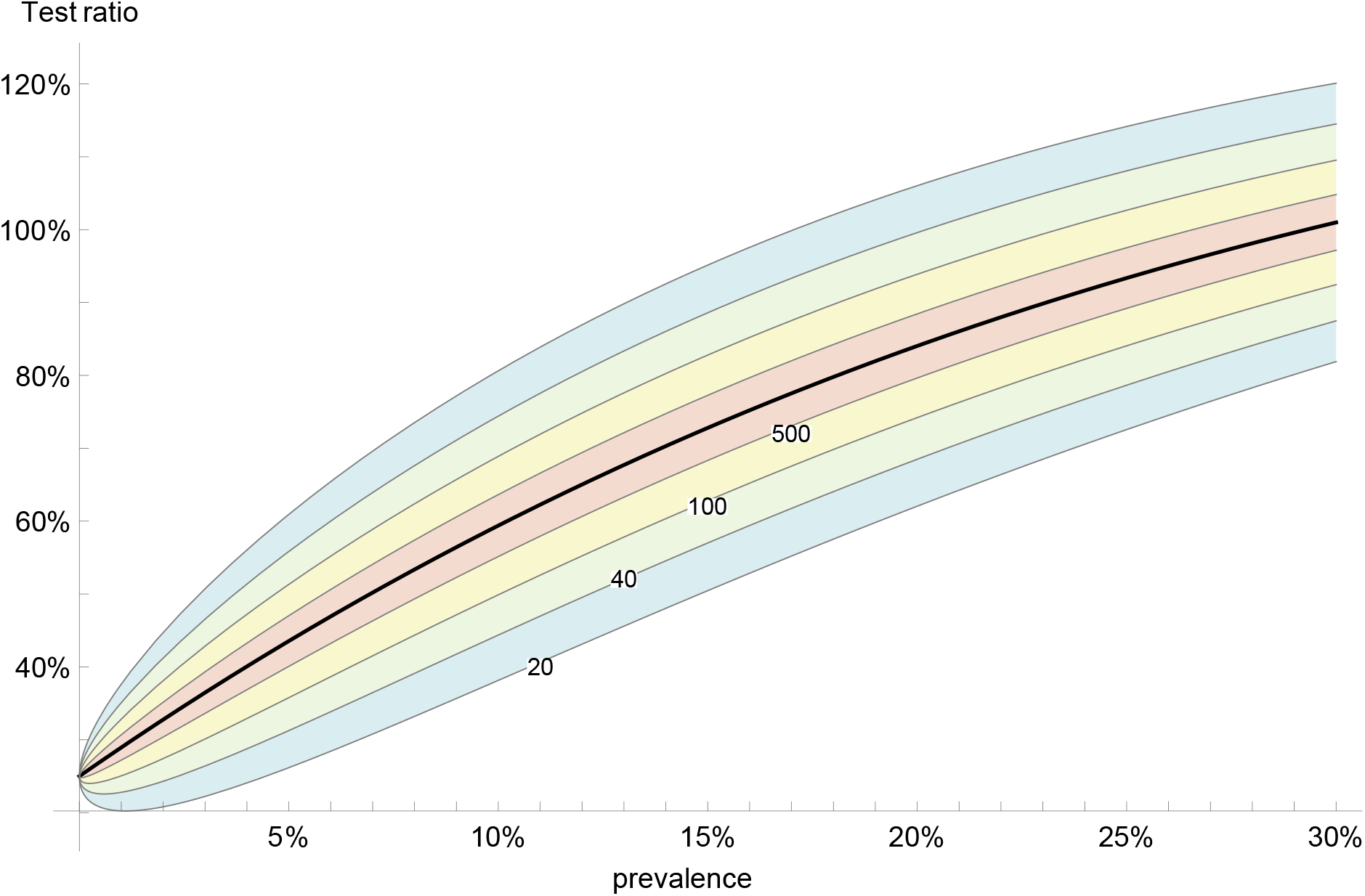
Expected test ratio with pooling (black curve) as function of prevalence, for pool size *M* = 4. The colored areas indicate the standard error (1*σ*), for different number of samples *N* = 20, 40, 100, 500.

The optimal pool size has been studied before and answer that has been given, e.g. [1, 4], is strongly dependent on the prevalence. This, however, is of limited usefulness because (i) the prevalence is unknown and (ii) it is unknown which statistical population is being sampled. Instead we propose to consider the prevalence itself to be a random variable, drawn from a uniform distribution. This amounts to sampling first from a population, and then sampling from a population of populations. Such a model may be not too far from reality. Consider, for example, one day a lab testing for Covid-19 all people from one workplace, another day prevalence testing travelers returning from a town, and so on. The choice of a uniform distribution for the prevalence suggests itself because nothing is known; such assumptions are known as the principle of indifference.

We average the expectation value of the test ratio *t* = 1*/M* + 1 *−* (1 *−p*)^*M*^ with respect to *p*, when *p* is uniformly distributed between *p*_1_ and *p*_2_. The resulting test ratio is: *⟨t⟩* = 1*/M* + 1 + [(1 *− p*_2_)^*M*+1^*−* (1 *−p*_1_)^*M*+1^]*/*[(*p*_2_*− p*_1_)(*M* + 1)]. The results in Figures 1, 2 are calculated from this expression.

The test errors can be calculated based on standard formulas for binary classification tests. We assume no dilution errors (see above for explanation). This means, for example, that the probability of obtaining a positive test result for one infected person is the same as for a pool of *M* people where at least one person is infected, even though the pool may contain a different amount of viral material.

Consider single-test sensitivity *s* and specificity *z*. The sensitivity and specificity of the pooled test be denoted by *s*′, *z*′ respectively. Following the two-stage protocol (as described above), one obtains: *s*′ = *s*^2^. For the specificity, one has the expression

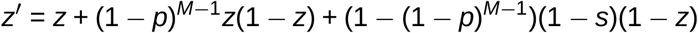

Here *M*≥2 is the pool size and *p* is the prevalence. The specificity of the pooled test is increased compared to the single-test specificity, as the right-hand side of the expression is always positive.

According to the double-average model, the expression for *z*′ can be averaged over a uniform distribution of prevalences. This gives

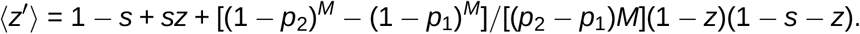

A plot, for some example values, is shown in Fig. 3.

## Conclusions

Pooling of tests allows to increase the test capacity for Covid-19 testing. For the classic two-stage protocol, a pool size of four is recommended, based on our model. In the absence of dilution errors, testing errors do not adversely affect the outcome.

## Data Availability

Article contains to external data.

## References

[1] Dorfman R. The Detection of Defective Members of Large Populations. The Annals of Mathematical Statistics. 1943;14(4):436–440. Available from: http://www.jstor.org/stable/2235930.

[2] Yelin I, Aharony N, Shaer Tamar E, Argoetti A, Messer E, Berenbaum D, et al. Evaluation of COVID-19 RT-qPCR test in multi-sample pools. Clinical Infectious Diseases. 2020 05. Available from: https://doi.org/10.1093/cid/ciaa531.

[3] Eis-Huebinger AM, Hoenemann M, Wenzel JJ, Berger A, Widera M, Schmidt B, et al. Ad hoc laboratory-based surveillance of SARS-CoV-2 by real-time RT-PCR using minipools of RNA prepared from routine respiratory samples. medRxiv. 2020. Available from: https://www.medrxiv.org/content/early/2020/03/31/2020.03.30.20043513.

[4] Abdalhamid B, Bilder CR, McCutchen EL, Hinrichs SH, Koepsell SA, Iwen PC. Assessment of Specimen Pooling to Conserve SARS CoV-2 Testing Resources. American Journal of Clinical Pathology. 2020 04;153(6):715–718. Available from: https://doi.org/10.1093/ajcp/aqaa064.

[5] Yuan yuan C. Statistical methods for batch screening of input populations by stage and group in COVID-19 nucleic acid testing. medRxiv. 2020. Available from: https://www.medrxiv.org/content/early/2020/04/28/2020.04.02.20050914.

[6] Farfan MJ, Torres JP, Oryan M, Olivares M, Gallardo P, Salas C. Optimizing RT-PCR detection of SARS-CoV-2 for developing countries using pool testing. medRxiv. 2020. Available from: https://www.medrxiv.org/content/early/2020/04/17/2020.04.15.20067199.

[7] Ben-Ami R, Klochendler A, Seidel M, Sido T, Gurel-Gurevich O, Yassour M, et al. Largescale implementation of pooled RNA-extraction and RT-PCR for SARS-CoV-2 detection. medRxiv. 2020. Available from: https://www.medrxiv.org/content/early/2020/05/07/2020.04.17.20069062.

[8] Torres I, Albert E, Navarro D. Pooling of Nasopharyngeal Swab Specimens for SARS-CoV-2 detection by RT-PCR. Journal of Medical Virology. 2020. Available from: https://onlinelibrary.wiley.com/doi/abs/10.1002/jmv.25971.

[9] Gupta E, Padhi A, Khodare A, Aggarwal R, Ramachandran K, Mehta V, et al. Pooled RNA sample reverse transcriptase real time PCR assay for SARS CoV-2 infection: a reliable, faster and economical method. medRxiv. 2020. Available from: https://www.medrxiv.org/content/early/2020/04/29/2020.04.25.20079095.

[10] Ghosh S, Rajwade A, Krishna S, Gopalkrishnan N, Schaus TE, Chakravarthy A, et al. Tapestry: A Single-Round Smart Pooling Technique for COVID-19 Testing. medRxiv. 2020. Available from: https://www.medrxiv.org/content/early/2020/05/02/2020.04.23.20077727.

[11] Schmidt M, Hoehl S, Berger A, Zeichhardt H, Hourfar K, Ciesek S, et al. FACT-Frankfurt adjusted COVID-19 testing-a novel method enables high-throughput SARS-CoV-2 screening without loss of sensitivity. medRxiv. 2020. Available from: https://www.medrxiv.org/content/early/2020/05/01/2020.04.28.20074187.

[12] Cleary B, Hay JA, Blumenstiel B, Gabriel S, Regev A, Mina MJ. Efficient prevalence estimation and infected sample identification with group testing for SARS-CoV-2. medRxiv. 2020. Available from: https://www.medrxiv.org/content/early/2020/05/06/2020.05.01.20086801.

[13] Becker MG, Taylor T, Kiazyk S, Cabiles DR, Meyers AFA, Sandstrom PA. Recommendations for sample pooling on the Cepheid GeneXpert® system using the Cepheid Xpert® Xpress SARS-CoV-2 assay. bioRxiv. 2020. Available from: https://www.biorxiv.org/content/early/2020/05/15/2020.05.14.097287.

[14] Hogan CA, Sahoo MK, Pinsky BA. Sample Pooling as a Strategy to Detect Community Transmission of SARS-CoV-2. JAMA. 2020 04. Available from: https://doi.org/10.1001/jama.2020.5445.

[15] Gan Y, Du L, Damola FO, Huang J, Xiao G, Lyu X. Sample Pooling as a Strategy of SARS-COV-2 Nucleic Acid Screening Increases the False-negative Rate. medRxiv. 2020. Available from: https://www.medrxiv.org/content/early/2020/05/26/2020.05.18.20106138.

[16] Lohse S, Pfuhl T, Berkó-Göttel B, Rissland J, Geißler T, Gärtner B, et al. Pooling of samples for testing for SARS-CoV-2 in asymptomatic people. The Lancet Infectious Diseases. 2020. Available from: https://doi.org/10.1016/S1473-3099(20)30362-5.

